# Personalizing dietary interventions by predicting individual vulnerability to glucose excursions

**DOI:** 10.1101/2024.08.07.24311591

**Authors:** Victoria Brügger, Tobias Kowatsch, Mia Jovanova

## Abstract

Elevated postprandial glucose levels pose a global epidemic and are crucial in cardiometabolic disease management and prevention. A major challenge is inter-individual variability, which limits the effectiveness of population-wide dietary interventions. To develop personalized interventions, it is critical to first predict a person’s vulnerability to postprandial glucose excursions—or elevated post-meal glucose relative to a personal baseline—with minimal burden. We examined the feasibility of personalized models to predict future glucose excursions in the daily lives of 69 Chinese adults with type-2 diabetes (*M* age=61.5; 50% women; 2’595 glucose observations). We developed machine learning models, trained on past individual context and meal-based observations, employing low-burden (continuous glucose monitoring) or additional high-burden (manual meal tracking) approaches. Personalized models predicted glucose excursions (F1-score: *M*=74%; median=78%), with some individuals being more predictable than others. The low burden-models performed better for those with consistent meal patterns and healthier glycemic profiles. Notably, no two individuals shared the same meal and context-based vulnerability predictors. This study is the first to predict individual vulnerability to glucose excursions among a sample of Chinese adults with type-2 diabetes. Findings can help personalize just-in-time-adaptive dietary interventions to unique vulnerability to glucose excursions in daily live, thereby helping improve diabetes management.

## Introduction

Cardiometabolic diseases are escalating worldwide—highlighting the need for better strategies to reduce their impact^1,2^. For example, type-2 diabetes accounts for more than 90% of the 537 millions of diabetes cases globally^3^, and is expected to double to at least 1.3 billion by 2050^1^. One key factor in the management of type-2 diabetes and related cardiometabolic diseases is postprandial glucose, which refers to blood glucose levels following meal intake^4,5^. More specifically, postprandial glucose excursions—defined here as increases in glucose following meals that exceed an individual’s baseline levels^6,7^—contribute to elevated HbA1c levels and pose an independent risk factor of cardiovascular disease^8^. Importantly, these glucose excursions are measurable and modifiable on a daily basis. Increased evidence suggests that dietary changes that reduce the intensity and frequency of glucose excursions can help manage and potentially prevent these diseases^9–12^.

Relatedly, diet is a crucial intervention target for glycemic excursions and type-2 diabetes management more broadly^1,2^. However, people interact with and respond to foods very differently, which poses a key challenge in dietary intervention development^13,14^. Individuals’ blood glucose levels vary widely in response to the same foods^13,14^, and people live in diverse socio-cultural and economic environments, with varying eating preferences and habits^15,16^. Altogether, this complex inter-individual variability limits the effectiveness of population-wide dietary recommendations for glycemic health^17^. In response, recent work (see example Refs.^13,18,19,20^) suggests moving beyond ‘one-size-fits-all’ dietary recommendations, and towards personalized strategies that account for individual factors. However, there exists no formal, established method to implement such personalization effectively^21^.

Clinical trials have begun to explore personalized dietary approaches through deep phenotyping^22^, which involves integrating detailed patient data such as microbiota profiles, food logs, blood tests, and clinical history^23,24^ to improve metabolic parameters. One trial, for instance, tailored dietary recommendations using deep phenotyping to predict postprandial glucose predictions. The tailored diet led to short-term improvements in postprandial glucose levels for both healthy and pre-diabetic participants^10^. In another trial, personalized food scores were created through deep phenotyping, which included analyzing microbe data, dietary patterns, glucose monitoring, cardiovascular risk, and medical history in a representative sample of U.S. adults. The 18-week program, which also offered nutrition and lifestyle education, successfully reduced waist circumference but did not affect triglyceride levels^25^. Additionally, a recent study among newly diagnosed type-2 diabetes patients combined medical and dietary history, microbiome analysis, food logging, and nutritionist consultations to develop personalized dietary recommendations. These tailored recommendations resulted in better metabolic outcomes over six months compared to a standard Mediterranean diet^23^. While these personalization approaches show potential, they are often costly and require multiple in-person visits and clinical tests, thus making them less feasible and accessible in individuals’ daily lives^26^. A key open question is how to develop dietary interventions dietary interventions for cardiometabolic disease prevention and management that are more scalable, lower-burden, and more precise at the individual level?

Advances in digital health technologies offer promising solutions for developing personalized dietary interventions that can be more easily integrated into individuals’ daily lives^27,28^—without laboratory tests and in-clinic visits. An increasingly popular approach in this direction, drawing on the health behavior change and reinforcement literature, involves just-in-time adaptive interventions (JITAIs)^29^. JITAIs continuously collect individualized data from mobile devices (e.g., wearables and smartphones) with an aim to trigger support at the right time when it is needed. JITAIs aim to recognize individualized patterns and to respond to vulnerable states—periods or triggers of increased susceptibility to signals like postprandial glucose excursions^29^. For example, when detecting a high vulnerability state to glucose excursions, JITAIs can deploy, and dynamically adapt, personalized smartphone notifications with meal and dietary recommendations, in quasi real-time^30^.

While JITAIs have only begun to appear in diabetes self-management^31,32^,their application for dietary interventions for glycemic control, particularly for type-2 diabetes, is still in its infancy. Thus, a crucial first step in developing dietary JITAI that aim to reduce the intensity and frequency of glucose deviations, is to identify factors that best predict an individual’s vulnerability to glucose excursions^4^. This involves pinpointing specific theory-driven triggers, such as meal content and contextual patterns^23^, that indicate when a person is most at risk for glucose excursions. For instance, analyzing an individual’s past meal habits and patterns, as well as temporal context, (e.g., mealtime) can help anticipate future vulnerability periods^33^. For some individuals, the intake of high-glycemic foods, like refined carbohydrates, may accurately predict glucose excursions^34^. For others, the main triggers could be the specific timing of their sweet intake after dinner; or a combination of mealtime and the nature of their past glucose excursions^35^. Understanding both the timing and composition of meals may be essential for making accurate, individualized predictions. However, the degree to which individuals differ in their vulnerability predictors has not been examined systematically, with some early studies often assuming that triggers (e.g., carbohydrate intake) remain consistent across individuals^36,37^. Mapping out individual vulnerability patterns to glucose excursions, may allow for new opportunities to derive key tailoring variables; meaning individual-level information that is used to decide when (i.e., under what conditions) to provide a dietary JITAI^29^.

To identify potential triggers of postprandial glucose excursions in individuals’ daily lives, two popular methods include manual meal tracking and continuous glucose monitoring (CGM). Meal tracking is often done through ecological momentary assessment (EMA), where individuals actively log their dietary intake and other behaviors using mobile apps^38,39^. While manual meal tracking can provide detailed information about meal content, it requires significant participant effort to log each meal, thus resulting in high data collection burden. On the other hand, CGM uses a small sensor placed on the arm to provide passive, near real-time monitoring of interstitial glucose levels, requiring no active user input^40^. Initially developed for type-1 diabetes care^41^, CGMs are increasingly used to manage type-2 diabetes, with increased reimbursement among individuals on insulin medication^40^. While less burdensome, CGMs are currently more costly than meal-tracking alternatives. Further, while CGM offers a low-burden, continuous monitoring of past glucose trends, manual meal logging provides more granular details about meal content but at the cost of higher user involvement.

Together, both data streams can be used to develop individual vulnerability profiles to glucose excursions in daily life, and outside the clinic. These signals can then be integrated to predict vulnerability states and trigger dietary JITAIs, for example delivered through ubiquitous smartphones. However, an open question is to what extent CGM, and manual meal logging approaches provide complementary value for predicting postprandial glucose excursions, or if one method may be more applicable for some individuals over others. Examining this question is important to guide methodological decisions, about when to employ CGM or manual meal tracking collection for participant profiling, as these data collection methods may be tailored to an individual’s specific circumstances in dietary JITAI development.

To this end, this study employs both low-burden (passive CGM monitoring) and high-burden (manual meal logging) approaches to predict individual postprandial glucose excursions and examine individual differences in vulnerability states. Motivated by prior work^10^, we test the feasibility of personalized machine learning models to predict individual vulnerability to postprandial glucose excursions. We examine: (1) to what extent do low-burden and high-burden personalized models predict postprandial glucose excursions at the individual level? (2) How does the performance of low-burden and high-burden models vary across individuals? (3) How do vulnerability states, or predictors of postprandial glucose excursions, vary across individuals?

To examine these research questions, we analyzed ∼12-day of CGM, contextual, and meal patterns data collected from 69 adults with type-2 diabetes in Shanghai, China; a highly relevant context as China has one of the highest diabetes prevalence rates globally^42^. We developed personalized, machine learning models, trained on individual past meals and context-based features from the past ∼6 days, to predict vulnerability to future postprandial glucose excursions, within an observational, daily life setting. Following prior work^6^, we classified glucose excursions, as values that surpass the average of the previous postprandial glucose level relative to individual baselines, and we use F1-score as primary metric of model evaluation^43^, as a combined metric of precision and recall that is not sensitive to class imbalance^44^. This approach aims to lay the groundwork for signals into developing more precise dietary JITAIs for postprandial glucose excursions, and more broadly type-2 diabetes management.

## Results

### Descriptives

On average, participants (*N*=69) showed a baseline glycated hemoglobin (HbA1c) values at 67.6 mmol/mol (*SD*=24.2), and fasting glucose at 158.54 mg/dl (59.45), both consistent with previously established type-2 diabetes criteria^45,46^. Participants reported an average of 3.37 (*SD*=0.68) meals daily, over the course of approximately 12.9 (*SD*=2.18) days. On average, participants reported consuming a total of 958g (*SD*=327.83) daily. The average daily intake included: 233.9g (*SD*=137.6) of staples, 146.3g (*SD*=66.6) of animal foods, 167.7g (*SD*=98.4) of vegetables, 30.1g (*SD*=58.6) of fruits, 8.5g (*SD*=11.9) of legumes and nuts, 86.9g (*SD*=13.72) of dairy, and 1.57g (*SD*=4.25) of sweets. The average participant showed a postprandial glucose value of 155.19 mg/dl (*SD*=33.29 mg/dl) and had 37.6 postprandial blood glucose observations (*SD*=10.5) available for analyses. Overall, 2’595 postprandial glucose observations were included in analyses. From these values, 1’099 glucose observations were classified as glucose excursions, resulting in an overall balanced class split of 41.7% (*SD*=12.4%) on average per individual. Most participants (*n*=65; 94%) reported taking hypoglycemia medications. For more details see Table S1 for demographic characteristics of the sample; Fig. S1 for Study flowchart and participant exclusion criteria.

### Personalized models predict individual postprandial glucose excursions

In line with our first research question, we examined the degree to which personalized models predict future postprandial glucose excursions at the individual level. Overall, the best personalized models— combining low and high-burden approaches—predicted future individual glucose excursions with a mean F1-score of 74% (median=78%, *SD*= 17%). The low-burden approach, developed by training the model on ∼6 days of past individual-level CGM observations, resulted in a mean F1-score of 72% (median=77%, *SD*=19%). The high-burden approach, developed by training the model on additional manual meal tracking over ∼6 days resulted in a mean F1-score of 71% (median=75%, *SD*=19%). See Fig. 1A for model density plots; and see Table S2, ‘Model performance metrics’ for precision, recall, and additional performance metrics.

**Fig 1.**
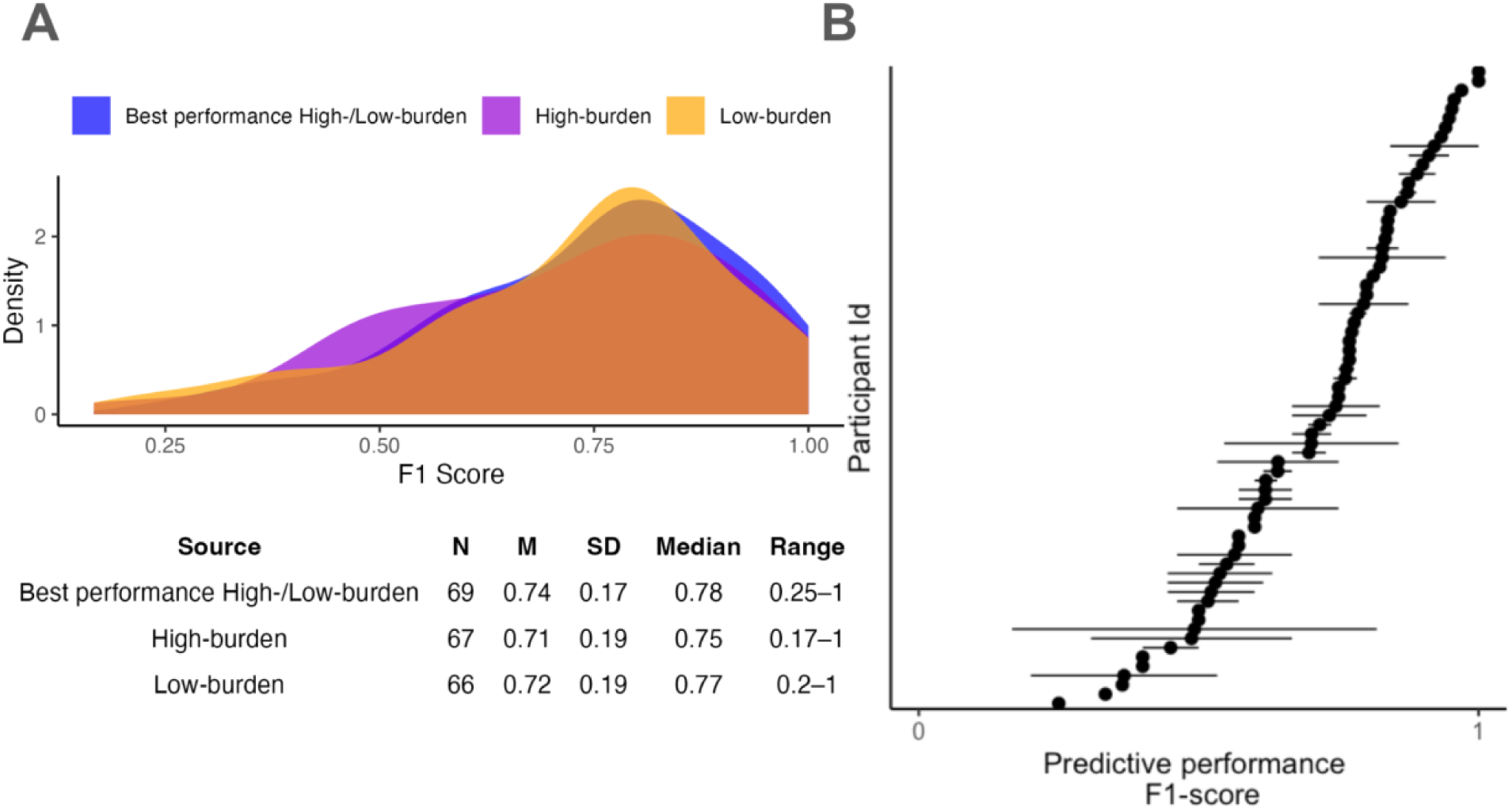
Model performance. (A) Performance metrics for the best personalized models, high-burden models, and low-burden models to predict individual postprandial glucose excursions. The best personalized model compares the performance of the low-burden and high-burden models and selects the higher-performing model for each individual. (B) Inter-individual variability in predictive performance across individuals, with dots showing the median across both low-burden and high-burden models per each individual; and the line showing the within-person range across both low-burden and high-burden model performance. Visualization for Fig. 1B is adapted from Refs.^47,48^ F1-scores are presented as decimal points.

Further, inspecting the F1-scores at the individual-level showed wide differences in predictability between individuals, and some degree of variability across data-collection approaches. As visualized in Fig. 1B, the range of F1-scores varied considerably between individuals, suggesting that postprandial glucose excursions were more easily predictable for some individuals than for others (overall range in F1-scores 17%-100%).

### Individual differences in low-burden vs. high-burden model performance

Our second research question examined individual differences in model performance across both high-burden and low-burden approaches—or the degree to which low-burden vs. high-burden models performed differently across individuals. No model performed the best across all participants. We found that the low-burden model performed better for 33% of the sample (n=23); the high burden model performed better for 28% (n=19) individuals; and no differences in model performance for the remaining 39% (n=27) participants. See Fig. 2 for individual slopes across both low- and high-burden models. Wilcoxon signed-rank tests showed that for the first subset, the low-burden model significantly improved the median F1-score by 12.2%, from 61.5% (high-burden model) to 73.7% (low-burden model) (Z =5.17, *p* <0.001, n=23; Fig. 2A). In contrast, for the second subset, the high-burden model significantly improved the F1-score median by 11.1%, from 66.7% (low-burden model) to 77.8% (high-burden model) (Z =3.80, *p* <0.001; n=19; Fig. 2B).

**Fig 2.**
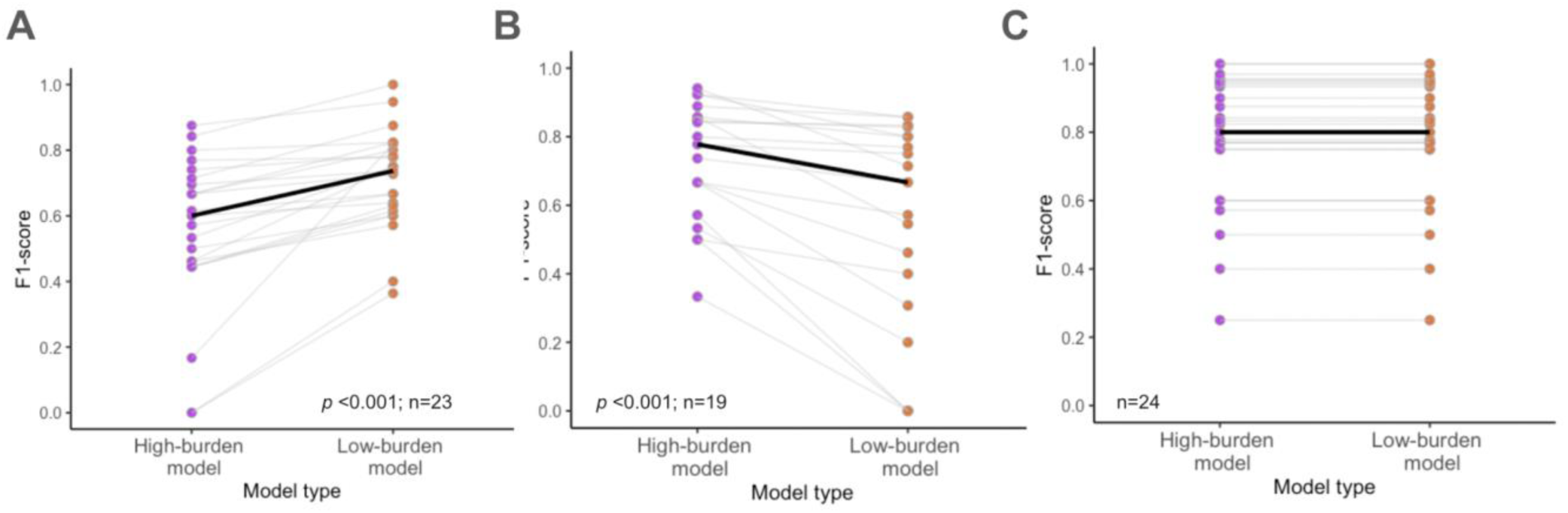
Individual differences in low-burden vs. high-burden model performance. F1-score comparison across both low-burden and high-burden models predicting individual postprandial glucose excursions. (A) Individuals for whom the low-burden (vs. high-burden) models performed better. (B) Individuals for whom the high-burden (vs. low burden) models performed. (C) Individuals for whom both low-burden and high-burden models performed similarly. Statistics are based on the paired Wilcoxon signed-rank test. Individual slopes are shown by gray lines, and the median is represented by the bold black line. Three individuals for whom the F1-scores could not be calculated were excluded from the visualization.

### Vulnerability predictors vary widely across individuals

We next examined which predictors (meal and temporal context-based) mattered most to predict future postprandial glucose excursions at the individual level, and how these “top” vulnerability predictors vary across individuals. Following Ref.^6^, we determined the percent importance of each feature from the total feature importance for the best performing model for each participant. See Table S3 for feature categorizations across additional high-burden and low-burden models.

As shown in Fig. 3, no two participants shared the same feature importance patterns, in both direction and strength. Individuals varied widely in which predictors best predicted future postprandial glucose excursions. For individuals for whom the high-burden model performed best, the top three features included total grams intake, vegetable intake, and the hour of day. However, neither of these top features were shared across all participants (See Fig. 3). The top feature, total grams intake, had importance scores ranging from 0% to 71.4% (*M*= 22.7%); and was only shared across 63% (*n*=12) participants. Vegetable intake had an average importance of 17.5%, ranging from 0% to 90.7%, across 63% (*n*=12) participants; and the hour of the day had an average importance of 9.5%, ranging from 0% to 51.9%, shared across 37% participants (n=7).

**Fig 3.**
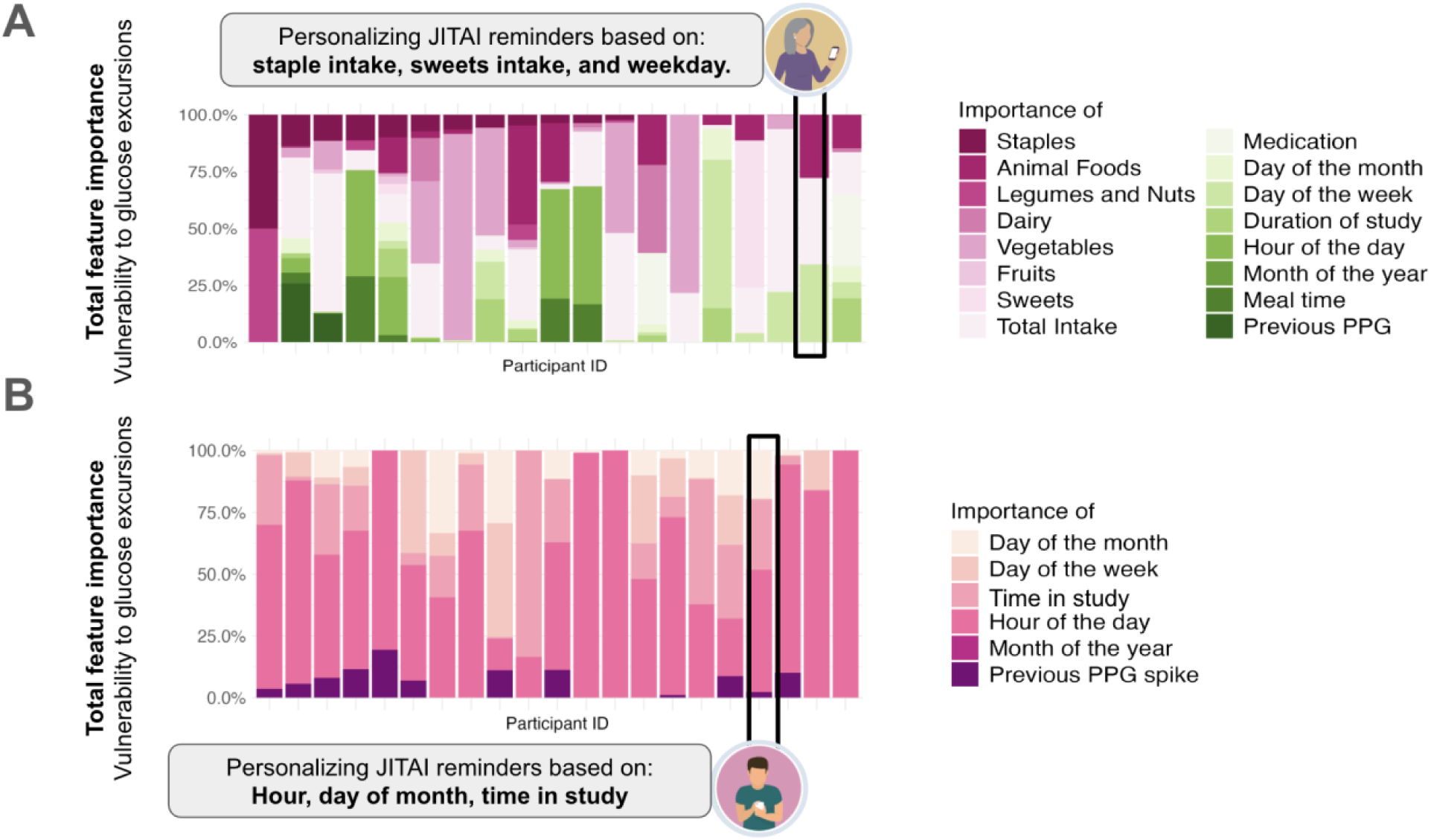
Feature importance predicting future postprandial glucose excursions. Importance among participants for whom (A) the high-burden model or the (B) low-burden model performed best, showcasing the diversity in the importance of predictors across individuals. The feature importance of each category (e.g., meals and temporal context) is represented by the percent importance from the total feature importance per individual^6^. Two individuals for whom feature importance could not be calculated were excluded in the low-burden model.

For individuals for whom the low-burden model performed best, the three top features included hour of day, time in study period, and day of the week. The top feature, hour of the day, had an average importance of 55.1% (range=0%-100%), shared across 91% (*n*=21) individuals. The day in the study period had an average importance of 16%, (range=0%-83.5%) and was shared across 70% participants (*n*=16). Further, day of the week had an average importance of 8.8%, (range=0%-46.2%), and was shared among 71% (*n*=15) individuals. See Fig. S2 and S3 additional feature importance plots for individuals for whom the high-burden and low-burden models performed equally, and for SHAP values^49^, showing the direction and strength of feature importance across high-burden and low-burden models.

### Personalizing dietary JITAIs based on individual vulnerability to postprandial glucose excursions

Mapping individual vulnerability patterns to postprandial glucose excursions can provide new opportunities to guide when (i.e., under what conditions) to provide a dietary JITAI^29^. For example, for the participant highlighted in Fig. 3A, postprandial glucose excursions were best predicted by their prior staple and sweet intake. For this individual, text-message notifications could be developed around the key mealtimes when staple and sweet intake combined is most expected. For example, on Friday evenings, when an individual is most likely to consume staples and sweets (based on past individual meal profiling), a notification could advise them to avoid these combinations and to instead opt for lower-glycemic alternatives. Here, lower-glycemic alternative recommendations could be derived based on meal contents that were most likely to reduce postprandial glucose excursions in the past. In turn, postprandial glucose excursions for the participant illustrated in Fig. 3B, are mainly predicted by temporal context-based factors (e.g., time of day). Here, time-sensitive notifications could be optimized during key risk periods, such as breakfast, identified as a highly vulnerable period for this individual. Drawing on behavior change theories, alternative breakfast choice recommendations could be paired with motivational messages such as cues to action from the Health Belief Model or self-efficacy messages from Social Cognitive Theory^50^ to motivate dietary changes. These two examples demonstrate how modeling inter-individual variability to glucose excursions can help design and trigger personalized JITAIs in near-real time. By integrating vulnerability triggers, dietary interventions may be more effectively customized to meet each person’s unique needs, thereby improving their postprandial glucose levels and overall diabetes management.

## Discussion

Individuals vary widely in their dietary patterns, habits, and daily environments, making it difficult to apply universal dietary recommendations for diabetes management effectively. This inter-individual variability underscores the need for personalized dietary intervention strategies^18,13,20^, particularly for managing postprandial glucose excursions in type-2 diabetes. In this study, we aimed to set the groundwork for dietary JITAIs by developing personalized machine learning models for predicting, and potentially preventing, postprandial glucose excursions, a key outcome in type-2 diabetes management^4^.

As a foundational step in JITAI development^51^, it is essential to first identify what factors best predict an individual’s vulnerability to postprandial glucose excursions. More specifically, we developed personalized machine learning models using two approaches: a low-burden method involving week-long CGM and a high-burden approach with additional manual, meal tracking to predict individual postprandial glucose excursions. We analyzed meal and context-based patterns among a sample of Chinese adults with type-2 diabetes, who were taking hypoglycemia medication. We aimed to evaluate model performance and to examine inter-individual variability in predictability and in feature importance across individuals.

Overall, we found that personalized models were able to predict future glucose excursions (F1-score: *M*= 74%; median=78%), similar to scores in prior work^10^. However, we observed substantial inter-individual variability. For some participants, the low-burden model was more predictive; for others, the high-burden model performed better, or showed no difference. Notably, the importance of meal and context-based features predicting postprandial glucose excursions varied widely across individuals. These results indicate that is no single vulnerability to glucose excursions shared across all participants.

These results extend prior work in several ways. First, earlier studies, (e.g. Refs.^10,52^) have highlighted the promise of deep phenotyping using lab data (e.g., combining blood tests, microbiome profiling, health history, nutrition consultations) to predict glycemic responses and tailor dietary interventions, however the trade-offs between comprehensive data collection and associated time, burden, and cost remain unclear. An open question is whether individual phenotyping to personalize dietary interventions could be completed with lower-burden alternatives in individuals’ daily lives. To this end, our study included both low-burden CGM and high-burden manual meal tracking approaches at the individual level, thus offering a comparison of their performance. Our results, suggest that ∼6 day remote participant profiling (using CGM or manual meal-tracking) may offer a more scalable substitute for predicting postprandial glucose excursions, and personalizing dietary interventions, relative to more burden intensive lab-based phenotyping profiling approaches^10^. However, future research may compare different profiling methods systematically, with further cost-benefit analysis.

Both CGM and manual meal tracking approaches can be integrated into JTAIs as individuals’ go about their daily lives^29^. However, our results suggest that these approaches may perform differently across different individuals to predict postprandial glucose excursions, which can inform decisions on when to select one approach over another. To explore which model worked better for whom, we conducted follow-up analyses to test if baseline characteristics, such as glycemic profiles (baseline HbA1c and fasting glucose), dietary patterns (based on manual meal logging), or demographics, were associated with the individual differences in low-versus high-burden model performance. Latent profile analysis indicated that individuals with lower BMI, healthier glycemic profiles (lower baseline HbA1c and fasting glucose), more consistent mealtimes over the meal logging period, and higher intake of vegetables and staple foods were more accurately predicted by the low-burden (CGM monitoring) approach compared to the high-burden (meal tracking) approach (*X2* (4, N =69) = 14.485, *p* < .01). These findings suggest that baseline heuristics, e.g., meal patterns and individual characteristics may perhaps help used to predict which approach is more predictive, and when, thus optimizing data collection. However, further research is necessary to validate these findings in a confirmatory setting.

While prior studies have developed several different algorithms for postprandial glucose prediction^6,10^, these have yet to be integrated into dietary JITAI frameworks. We extend this line of work by highlighting the feasibility of short-term (∼6 day) CGM and meal-tracking participant profiling to capture individual vulnerability to postprandial glucose excursions in a type-2 diabetes context, and to provide time-sensitive tailoring variables for JITAI development. However, to fully integrate these predictive models, future research should move from observational studies to controlled experiments; thus, validating these personalized models into a micro-randomized JITAI framework and comparing their effectiveness to generic, population-wide dietary interventions.

Our study offers high ecological validity by being conducted in a real-life setting, which enhances the relevance of our findings. Further, this study is the first to our knowledge to predict individual vulnerability to postprandial glucose excursions among a sample of Chinese adults with type-2 diabetes; with implications to integrate these individual differences into JITAI development. However, several limitations should be considered. Firstly, the dietary logs focused on meals, which may under report snacking. For example, participants reported very low consumption of sweets during the tracked period. Additionally, the sample comprised primarily Chinese adults, who reside in Shanghai with type-2 diabetes who were predominantly using hypoglycemia medications, which may limit the generalizability of the results^53^. For example, individuals with different ethnicities vary in baseline postprandial glucose levels^54^. To broaden the generalizability of these findings, further research is necessary with diverse ethnic groups, non-medicated individuals, and populations with type-1 diabetes or prediabetes. Future studies should also consider incorporating automated food tracking technologies to reduce participant burden of manual food logging^55^. Further, incorporating additional wearables to monitor sleep, physical activity, heart rate variability, and stress, can potentially enhance the predictability of postprandial glucose levels^56^. For example, research integrating wearable data has demonstrated that it is possible to improve the F1-score for postprandial glucose level predictions to 84-86% among healthy individuals^6^. Nevertheless, our current findings suggest that CGM and temporal data alone may already provide a promising level of predictability, for some individuals in the type-2 diabetes context.

In conclusion, our findings show that personalized machine learning models, drawing on CGM or manual meal tracking, can feasibly predict future postprandial glucose excursions among a sample of Chinese adults with type-2 diabetes, with model performance and predictors varying considerably among individuals. By employing ∼6-day CGM or manual-meal tracking profiling, interventionists can identify individual vulnerabilities to glucose excursions, thus laying the groundwork for dietary JITAIs. Future research should integrate these predictive models into JITAIs to further optimize diabetes care.

## Methods

We used a publicly available, observational dataset focusing on individuals with type-2 diabetes recruited in Shanghai, China ^57^. The dataset is made public with an aim to advance machine learning models to optimize personalized lifestyle recommendations and glycemic control. In this dataset, patients were recruited from Diabetes Data Registry and Individualized Lifestyle Intervention in Shanghai East Hospital (September 2019 to March 2021) and Shanghai Fourth Peopleś Hospital (June 2021 to November 2021), respectively. As we only used anonymized data, ethics approval was not applicable. The original data collection study was approved by the Ethics Committee of Shanghai Fourth People’s Hospital and Shanghai East Hospital affiliated to Tongji University in accordance with the Declaration of Helsinki. Informed consent was obtained from all the patients.

### Participants

Eligible participants included 106 patients diagnosed with diabetes-type 2 according to the 1999 World Health Organization (WHO) criteria ^58^, aged 18 or older, willing to sign the informed consent form, and having CGM recordings for at least 3 days. Patients were excluded if they reported alcohol or drug abuse, were unable to comply with the study, or were deemed unsuitable by the original investigators who completed the data collection. Our analysis included 69 participants with type-2 diabetes, with a total 2’595 postprandial glucose observations. Thirty-seven participants were excluded from our analyses: *n*=1 for having no postprandial glucose measures, *n*=36 for not having insufficient postprandial glucose measures to split the data in a train-test split (<20 observations in total), and *n*=3 due to insufficient variability to run the predictive models, with all observations belonging to the same class (see Supplement Flowchart Fig S1 ‘Study flowchart and exclusion criteria’).

### Study procedure

As part of the data collection, participants were instructed to wear a continuous glucose monitoring device (FreeStyle Libre H, Abbott Diabetes Care, Witney, UK) to measure interstitial glucose levels continuously for up to 14 days, and to report the contents of their meals, and their diabetes medicine intake using event-based EMA surveys. Participants also provided laboratory blood measurements, collected 6 months prior to the study.

### Measures

Data for model building included EMA-based meal content records, diabetes medicine intake records, CGM recordings, and temporal, context-based measures. See Supplement Tab. S3 ‘Features categorization’ for the full feature list and categorization.

#### Meal content features

Participants logged their meals throughout the study, and these logs were categorized into food groups commonly consumed in China^59^, which include: (1) staples (e.g., rice, noodles), (2) vegetables, (3) fruits, (4) animal foods (e.g., chicken, eggs), (5) dairy products, (6) legumes, nuts, and seeds, and expanded with (7) sweets, which are associated with blood glucose levels. These food groups were used once as binary variables and once as food group grams per meal. Additionally, total grams of food intake were assessed. All these variables were calculated for the current period and lagged for the previous 8-hour intake and the previous 24-hour intake based on prior work. Resulting into 45 dietary features.

#### Diabetes medicine-intake features

Participants recorded their daily insulin and non-insulin diabetes hypoglycemia medication intake including the exact timing. Medication logs were converted into time-varying variables to account for the different effects and doses of diabetes insulin medications on blood sugar levels over time. Basal and intermediate-acting insulins have a gradual effect over 2 to 24 hours ^60,61^; bolus insulin acts quickly within 15 minutes to 4 hours^62^; and non-insulin hypoglycemic agents start working immediately and last for about 6 hours^63^. Leading to 10 diabetes medicine-intake features.

#### Temporal features

Temporal features were created from the time stamps in the EMA logs. Variables were created for meal time, day of the month, months of the year, hour of the day and day of the study duration. To account for possible fasting periods, binary meal time indication in the last 8 hours and 24 hours, day of the week, were also included.

### Postprandial glucose excursions

Based on previous research^6^, we calculated personalized postprandial glucose excursions, relative to an individuals’ baseline, as our main outcome. A binary variable for 2-hour postprandial glucose was determined after each self-reported meal, indicating whether it was higher or lower than the previous mean postprandial glucose measure, thus providing each participant with a personal rolling baseline. This allowed us to compute postprandial blood glucose excursions approximately 2 hours after each meal relative to each participant’s previous value. The first observation was excluded as no previous mean postprandial glucose could be determined. We also included each individual’s lagged value as a feature to reflect their postprandial glucose levels from the meal before.

### Data analyses

#### Model development

We developed two personalized machine learning models for each participant, a low-burden and a high-burden model. The low-burden model included the following features: previous postprandial glucose excursion, medicine-intake (non-insulin hypoglycemic agents and bolus, basal, and intermediate insulin intake, dose, and effect time), and temporal features (such as mealtime, time of day). The high burden-model included the same features, in addition to dietary intake (food groups and total intake per meal, in the last 8 and 24 hours). See Supplement Table S3 ‘Features categorization’ for full features and their categorization. Any missing values were imputed using single imputation, and missingness dummy variable columns were introduced in the model.

To predict postprandial glucose excursions, we employed personalized Extreme Gradient Boosting (XGBoost) ^64^, a composite learning algorithm used in similar, prior work^65^, and used F1-score as our main model performance metric. All analyses were conducted using R version 4.3.2.

#### Rolling origin cross-validation

For each individual, we employed a 70/30 within-person training-test split due to the time series nature of the data. The first 70% of the observations served as the training set, while the remaining 30% was used as the test set. To determine the final model, we performed time series rolling origin cross-validation (CV) with four folds on the training set. Rolling origin cross-validation involves sequentially expanding the training data window while keeping the test data fixed. This approach is specific to time-series data and ensures that the model does not use future information to predict past events^48,66^.

#### Hyper-parameter tuning

Hyperparameters for XGBoost were determined using a 4-fold cross-validation (CV) grid search method (nrounds = seq(from = 50, to = 150, by = 50); eta = c(0, 0.0001, 0.001, 0.01, 0.015); max_depth = c(1, 2, 3); gamma = c(−0.5, 0, 0.5); colsample_bytree = c(0.5, 1, 2); min_child_weight = c(1, 2); subsample = 1), which systematically explores various combinations to find the optimal settings based on predefined criteria.

#### Feature importance

To determine the feature importances for both high-burden and low-burden models, we used eXtreme Gradient Boosting (XGBoost) importance and SHAP value per participant^49^.

## Data Availability

All data produced are available online at:

https://figshare.com/collections/Diabetes_Datasets_ShanghaiT1DM_and_ShanghaiT2DM/6310860/2

